# Circulating CTHRC1 Levels are Associated with IPF Disease Severity and Survival

**DOI:** 10.1101/2025.11.24.25340889

**Authors:** Monica M. Yang, Tatsuya Tsukui, Michael Wax, Seoyeon Lee, Averey Lea, Alexey Bazarov, Lisa Hazelwood, Paul J. Wolters, Dean Sheppard

**Affiliations:** Department of Medicine, Division of Rheumatology; Pulmonary, Critical Care, Allergy and Sleep Medicine, University of California, San Francisco, CA, USA; AbbVie, Inc, North Chicago IL, USA

## Abstract

**Objective:** Idiopathic pulmonary fibrosis (IPF) is a disease of high morbidity and mortality. We previously identified a novel subset of pathologic fibroblasts, characterized by CTHRC1 expression, uniquely present in fibrotic lung diseases. The aim of this study was to determine the association of serum CTHRC1 with clinical outcomes in IPF to assess its potential as a biologically relevant biomarker.

**Methods:** A retrospective longitudinal cohort study was performed utilizing two cohorts including a discovery cohort of 352 IPF patients from University of California San Francisco (UCSF) and a validation cohort of 1,152 IPF patients from the Pulmonary Fibrosis Foundation (PFF) as well as 41 healthy controls. Serum CTHRC1 was measured by ELISA and patients were stratified by quartiles of CTHRC1 for analysis. For a subset of patients, serial serum and lung CTHRC1 expression were measured. Associations between CTHRC1 and clinical outcomes, including baseline lung function, lung function trajectory, and transplant-free survival were assessed.

**Results:** Serum CTHRC1 was elevated in IPF patients compared to healthy controls (UCSF 31661±11651 and PFF 33916±14547 vs. 24409±8630pg/ml, p<0.001). Elevated circulating CTRHC1 was associated with lower FVC% and DLCO% at baseline and a greater decline in FVC over one year. In both cohorts, higher CTHRC1 level was associated with worse transplant-free survival (p<0.03).

**Conclusion:** Circulating CTHRC1 levels are elevated in patients with IPF and associated with disease severity and overall survival. These findings further support the biological significance of CTHRC1 in IPF as a pathologic marker and potential biomarker reflecting the burden of pathologic fibroblasts in IPF.

**Key Messages:** What is already known on this topic:

IPF is a progressive fibrotic lung disease driven in part by activation and expansion of pathologic fibroblasts.

CTHRC1+ fibroblasts have been identified in fibrotic lung tissue of IPF and are implicated in fibrogenesis.

While CTHRC1+ expression in lung tissue has been established, its role as a potential circulating biomarker has yet to be studied.

What this study adds:

This study demonstrates serum CTHRC1 levels are significantly elevated in patients with IPF compared to healthy controls.

Elevated serum CTHRC1 levels are associated with decreased baseline lung function, greater disease progression, and reduced 5-year transplant-free survival.

How this study might affect research, practice or policy:

These findings support CTHRC1 as a biologically relevant biomarker for prognostication in IPF.

## Introduction

Idiopathic pulmonary fibrosis (IPF) is a chronic lung disease characterized by epithelial cell failure and excess matrix deposition of the lung parenchyma, leading to lung scarring and progressive respiratory decline.^1^ IPF is the most common form of interstitial lung disease and is associated with significant morbidity and mortality, with a median survival of three years following diagnosis.^2,3^ While mortality is high, the clinical course of IPF may vary between individuals ranging from slow to rapidly progressive disease.^4^ This clinical heterogeneity has made management of the disease difficult as there are limited prognostic markers that predict outcomes, stratify patients, or guide treatment. Identifying clinically actionable biomarkers is needed to advance patient care and precision medicine in the field.

IPF is a disease of epithelial dysfunction that leads to pathologic fibroblast activation, differentiation, and proliferation.^5,6^ Pathologic fibroblasts are responsible for excessive extracellular matrix (ECM) deposition, which disrupts normal lung architecture, impairing oxygen exchange. The advent of single-cell RNA sequencing (scRNA-Seq) has enabled the detailed characterization of pathologic cell types within the fibrotic lung, allowing for identification of novel targets and markers of interest.^7–9^ We identified a novel subset of pathologic fibroblasts characterized by CTHRC1 expression that are uniquely present in fibrotic lung diseases.^10^ These CTHRC1+ fibroblasts were shown to express the highest levels of ECM proteins, are localized predominantly in fibroblast foci, and have increased migratory capacity, supporting their profibrotic phenotype. Lineage tracing further showed these CTHRC1+ fibroblasts to emerge from alveolar fibroblasts after lung injury to propagate fibrogenesis.^11^

CTHRC1 is a secreted glycoprotein that has potential to serve as a biologically relevant circulating biomarker representing the burden of profibrotic fibroblasts in IPF.^12^ While its exact function is unknown, CTHRC1 has been implicated in cell migration and tissue remodeling after injury, and CTHRC1 serum levels have been found to be elevated in diseases including rheumatoid arthritis, hepatic fibrosis, and several malignancies.^13–17^ Although expression of CTHRC1 in pathologic fibroblasts is now well established in IPF, the importance of circulating CTHRC1 in IPF has yet to be explored.^10,11,18^ The objective of this study was to further understand the pathologic relevance of CTHRC1 by exploring the relationship between circulating CTHRC1 levels with pulmonary outcomes in IPF and assess its utility as a prognostic biomarker.

## Methods

### CTHRC1 expression in IPF lung

ScRNAseq data were acquired from human lung tissue as previously described (GSE132771).^10^ The data was subsetted to include IPF and Healthy Control subjects and analyzed using Seurat 5.1.0 and visualized using scCustomize 2.1.2.^19,20^ Following unsupervised clustering, cell annotation was performed using canonical markers as described previously. CTHRC1 gene expression was compared between IPF and healthy control lung in all cells as well as individual cell types using Wilcoxon Rank Sum test.

### Study Populations

This is a retrospective longitudinal cohort study utilizing two cohorts including a discovery cohort of 352 IPF patients from the University of California San Francisco (UCSF) Interstitial Lung Disease Patient Registry and Biorepository and a validation cohort comprising 1,152 IPF patients from the Pulmonary Fibrosis Foundation (PFF) Patient Registry, a multicenter observational cohort.^21^ All patients met the diagnostic criteria for IPF based on ATS/ERS/JRS/ALAT clinical practice guidelines.^22^ 41 healthy controls from UCSF were also included in this study. This study was approved by the Institutional Review Board at UCSF and written informed consent was obtained from all subjects.

### Clinical Data and Endpoints

Demographic and clinical data including age, sex, race, smoking status, and antifibrotic use were recorded at time of baseline blood draw. Additional clinical data including relevant co-morbidities, family history of ILD, and disease duration were recorded in the UCSF cohort.

Pulmonary function test (PFT) data was obtained at baseline and abstracted at 12-month intervals, within a 3-month time frame, for 48 months. The primary outcome of interest was forced vital capacity percent predicted (FVC%) with diffusing capacity of carbon monoxide percent predicted (DLCO%) and transplant free survival as secondary outcomes. Survival was recorded within 5 years of baseline with death and lung transplantation considered terminal events.

### CTHRC1 measurement in serum

Baseline serum samples were collected at time of cohort enrollment and stored at -80C prior to analysis. Sera were diluted 1:20 in assay diluent and circulating CTHRC1 levels were measured by commercial ELISA according to the manufacturer’s protocol (Abcam 274399, Abcam Inc. Waltham, MA). All samples were run in duplicate and mean CTHRC1 levels were used in analyses.

### Statistical Analysis

Statistical analysis was performed using R version 4.4.0. Patient characteristics were summarized by mean and standard deviation for continuous variables and frequency for categorical variables. Association of circulating CTHRC1 level with clinical variables was compared using Wilcoxon and Kruskal Wallace. Multivariable linear regression analysis was performed to identify associations between clinical factors including age, sex, race, smoking, comorbidities, disease duration, and antifibrotic use with CTHRC1 level. Multicollinearity among predictors was assessed using the variance inflation factor (VIF), with VIF values > 5 indicative of possible collinearity. For additional analysis, patients were stratified by quartiles of CTHRC1 level with Q1 having the lowest CTHRC1 level and Q4 having the highest. Association between CTHRC1 quartile and clinical outcomes was evaluated using Wilcoxon and multivariable modeling was used to account for potential confounders. Survival analysis of 5-year transplant-free survival, defined as time to death or lung transplantation, was performed using Kaplan-Meier curve and log-rank test for significance. Survival time was censored when a patient was lost to follow-up within the 5-year period. Cox proportional hazard model adjusting for age, sex, smoking status, and baseline FVC% was used for hazard ratio estimation with the lowest quartile (Q1) serving as the reference group.

### Longitudinal CTHRC1 measurement

For a subset of patients with available serial serum samples, serum CTHRC1 was measured at a second time point after baseline using the method described above. To limit plate-plate variability, baseline and serial CTHRC1 samples were run on the same ELISA plate. Utilizing CTHRC1 at both timepoints and years between serum sample collection, a rate of change per year was calculated. Association of rate of CTHRC1 change with clinical variables was compared using Wilcoxon and Kruskal Wallace.

### CTHRC1 Measurement in Tissue with qPCR

To examine CTHRC1 levels at the organ level and its association with serum levels, CTHRC1 expression were measured in lung tissue using quantitative real-time PCR (qPCR). Total RNA was extracted using the RNeasy Mini Kit (Qiagen) according to the manufacturer’s instructions, and 1 µg of RNA was reverse transcribed using the SuperScript IV VILO Master Mix with ezDNase (Invitrogen). qPCR was performed using SensiFAST SYBR lo-ROX kit (Meridian Bioscience) on a QuantStudio 7 Pro Real-Time PCR System (Applied Biosystems). The amplification program was 95°C for 2 min followed by 40 cycles of combination of 95°C for 15 sec, 60°C for 30 sec and 72°C for 1min. Relative mRNA levels of target genes were normalized to GAPDH mRNA expression and analyzed by 2-ΔΔct method. The sequences of primers are listed in Supp 1.

## Results

### CTHRC1 expression is unique to pathologic fibroblasts in IPF lung

To evaluate the specificity of CTHRC1 expression for pathologic fibroblasts in IPF, we reanalyzed single-cell data from 3 IPF and 3 healthy control lungs (Figure 1A). Globally, CTHRC1 expression was increased in IPF and largely absent in healthy lungs (log2FC 4.16, p<0.005) (Figure 1B-C). Among cell subtypes, CTHRC1 was significantly increased in pathologic fibroblasts, compared to all other cells (log2FC 3.56, p <0.005). These pathologic fibroblasts were almost exclusively found in IPF lungs with a subset of them demonstrating the highest levels of CTHRC1 expression (Figure 1D). CTHRC1 was detected in other mesenchymal cell subtypes of healthy and IPF lungs, including pericytes and endothelial cells, however, at significantly lower levels. These data indicate that pathologic fibroblasts are the major source of CTHRC1 in IPF lungs.

**Figure 1:**
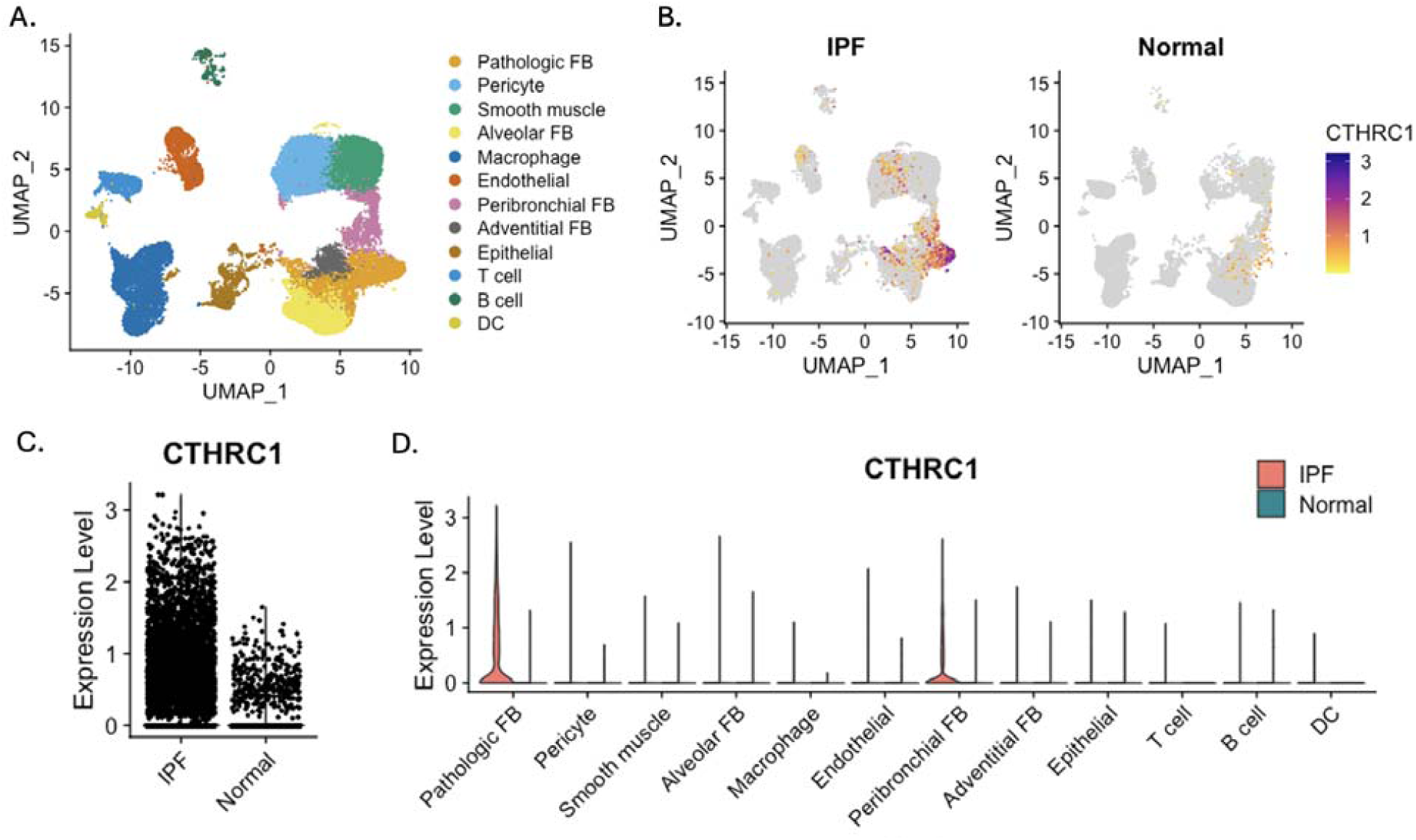
CTHRC1 expression is increased in IPF lung preferentially in pathologic fibroblasts (A) Uniform manifold approximation and projection (UMAP) plot of all cells from 3 IPF and 3 healthy control lungs. CTHRC1 expression by UMAP (B) and Violin Plot (C) by disease type demonstrating CTHRC1 is increased in IPF. (D) Violin plot comparing CTHRC1 expression by cell type between IPF and healthy controls, showing expression primarily in pathologic fibroblasts.

### Cohort Characteristics

The demographic data and clinical characteristics of 352 UCSF IPF patients, 1156 PFF IPF patients, and 41 healthy controls are shown in Table 1. Both UCSF and PFF Patient Registry cohorts were predominantly male and white with a prior smoking history. Mean disease duration of the UCSF cohort was 0.16 years with 90% of participants being enrolled at time of IPF diagnosis. There were 39 cases of familial IPF in the UCSF cohort and the most common comorbidities included gastroesophageal reflux disease (37%), coronary artery disease (24%), and chronic obstructive pulmonary disease (23%) (Supp 2). Comparing the UCSF and PFF cohorts, there were significant demographic differences. Notably, the PFF Patient Registry was more likely to be white and female, had a higher rate of anti-fibrotic use and lower baseline FVC% and DLCO% than the UCSF cohort. Lung transplantation rates were similar between the two cohorts.

**Table 1:** Demographic and Clinical Characteristics of Study Cohorts.

|  |  | IPF |  |  |
| --- | --- | --- | --- | --- |
|  | Normal<br>(N=41) | UCSF<br>(N=352) | PFF<br>(N=1156) | P-value |
| Age |  |  |  |  |
| Mean (SD) | 60.4 (7.13) | 70.3 (8.62) | 67.5 (10.6) | <0.001 |
| Sex |  |  |  |  |
| Female | 23 (56.1%) | 87 (24.7%) | 458 (39.6%) | <0.001 |
| Male | 18 (43.9%) | 265 (75.3%) | 698 (60.4%) |  |
| Race |  |  |  |  |
| White | 31 (75.6%) | 274 (77.8%) | 965 (83.5%) | <0.001 |
| Hispanic | 4 (9.8%) | 40 (11.4%) | 76 (6.6%) |  |
| Black | 0 (0%) | 2 (0.6%) | 61 (5.3%) |  |
| Asian | 6 (14.6%) | 29 (8.2%) | 34 (2.9%) |  |
| Other | 0 (0%) | 7 (2.0%) | 4 (0.3%) |  |
| Smoking |  |  |  |  |
| Yes |  | 230 (65.3%) | 668 (57.8%) | 0.0136 |
| No |  | 122 (34.7%) | 488 (42.2%) |  |
| Antifibrotics |  |  |  |  |
| Yes |  | 43 (12.2%) | 402 (34.8%) | <0.001 |
| No |  | 308 (87.5%) | 722 (62.5%) |  |
| Unknown |  | 1 (0.3%) | 32 (2.8%) |  |
| FVC, %predicted |  |  |  |  |
| Mean (SD) |  | 72.4 (18.1) | 67.1 (18.1) | <0.001 |
| DLCO, %predicted |  |  |  |  |
| Mean (SD) |  | 50.6 (17.5) | 42.2 (16.5) | <0.001 |
| Lung Transplant |  |  |  |  |
| Yes |  | 54 (15.3%) | 173 (15.0%) | 0.93 |
| No |  | 298 (84.7%) | 983 (85.0%) |  |

\*p-value calculated between UCSF and IPF cohorts. FVC, force vital capacity; DLCO, diffusing capacity for carbon monoxide; UCSF, University of California San Francisco Cohort; PFF, Pulmonary Fibrosis Foundation Patient Registry

### Serum CTHRC1 levels are elevated in IPF and associated with lower baseline FVC and DLCO

Patients with IPF had significantly higher serum CTHRC1 levels than healthy controls in both the UCSF and PFF (31661±11651 and 33916±14547 pg/ml vs. 24409±8630 pg/ml, p <0.001). Differences in CTHRC1 level were not observed between UCSF and PFF Patient Registry cohorts (Figure 2). In multivariable linear regression, age was significantly associated with CTHRC1 levels (β=276.2, p <0.001) with no association found between CTHRC1 levels and race, smoking, disease duration, comorbidities, family history of ILD or antifibrotic use. All predictors showed low multicollinearity (VIF < 1.25).

**Figure 2:**
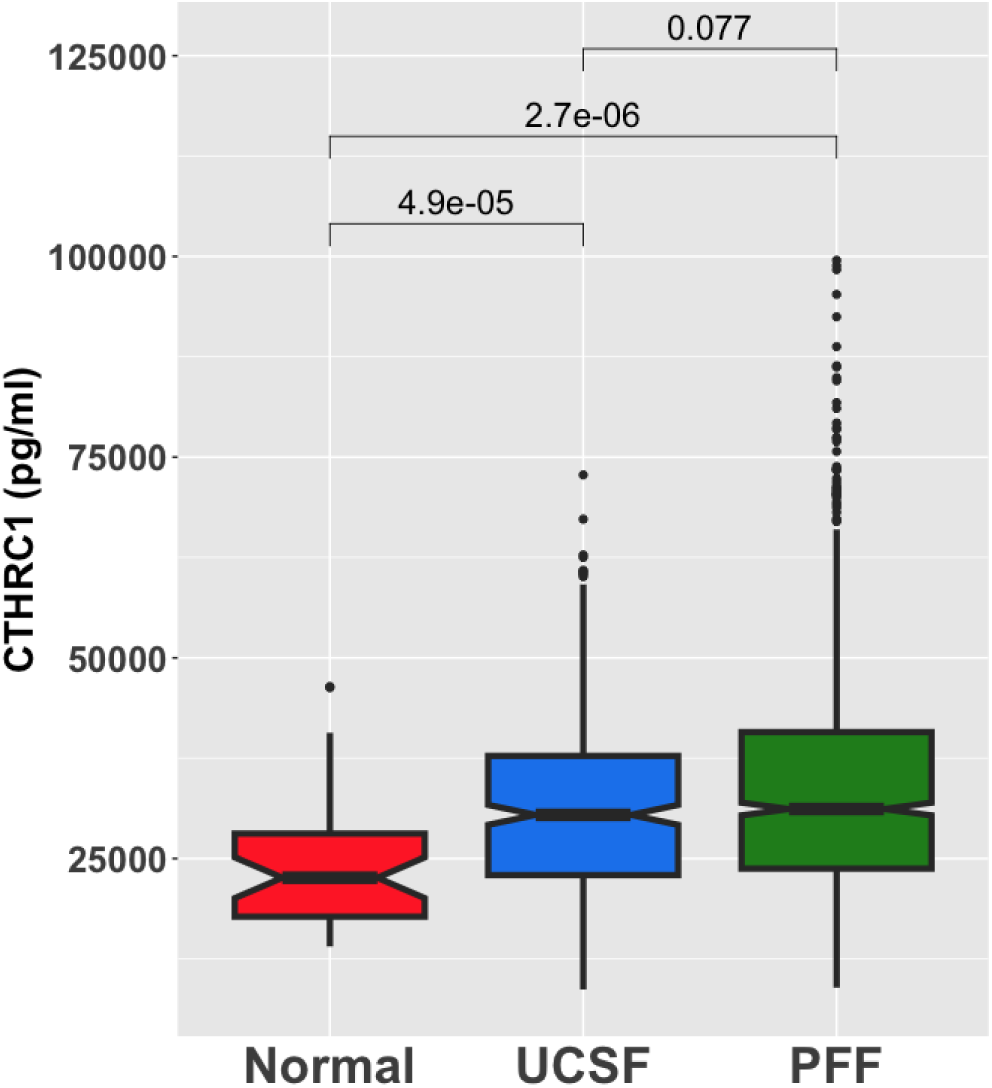
Serum CTHRC1 levels are elevated in IPF patients compared to controls. Notched boxplots of serum CTHRC1 level by patient cohort. Group comparisons measured by Wilcoxon Rank Sum test.

For subsequent analyses, cohorts were stratified into equal quartiles by CTHRC1 serum level (Q1 = lowest CTHRC1, Q4 = highest CTHRC1). After controlling for covariates, elevated CTHRC1 was significantly associated with lower FVC% in both the UCSF and PFF Patient Registry with a mean reduction in FVC% of 4.5% per quartile increase in the UCSF cohort (p=0.00012) and 3.2% in PFF (p = 0.004) (Figure 3). Similarly, higher CTHRC1 levels were also associated with lower DLCO% in both patient cohorts (mean DLCO% decrease per quartile increase 3.2% in UCSF p=0.029, 2.3% in PFF p = 0.0063).

**Figure 3:**
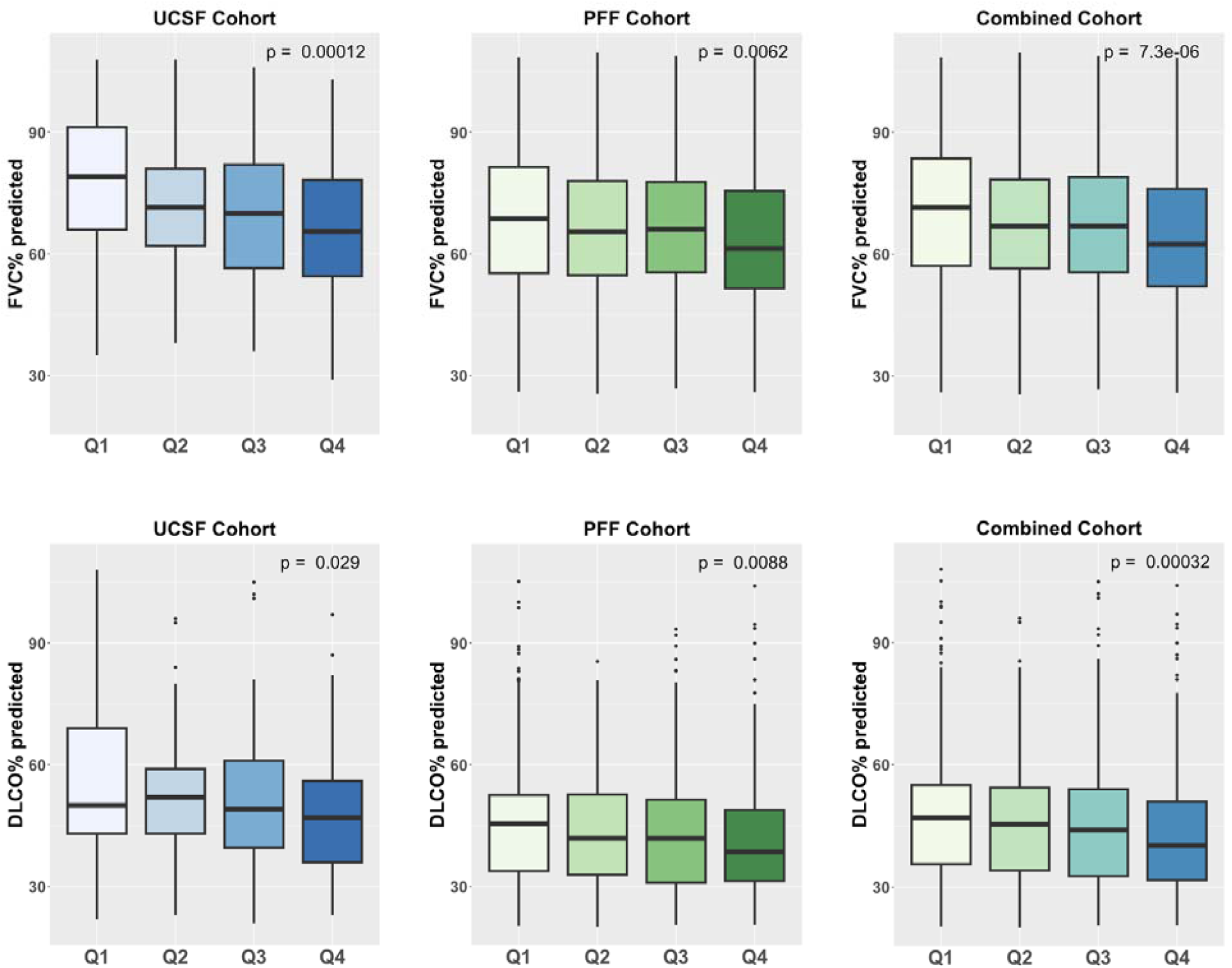
Serum CTHRC1 levels are associated with lung disease severity at baseline in IPF patients. Association of serum CTHRC1 levels in IPF with lung disease severity by patient cohort. Top: Comparison of FVC% between quartiles of serum CTHRC1 level in UCSF (left), PFF (middle), and combined (right) cohorts. Bottom: Comparison of DLCO% between quartiles among cohorts. Q1 = Lowest CTHRC1, Q4 = Highest CTHRC1.

### Serum CTHRC1 levels are associated with disease progression in IPF

To assess the association of serum CTHRC1 levels with disease progression, we examined change in FVC (ml) and DLCO% predicted over 1 year (Figure 4). A total of 1070 patients (279 UCSF, 791 PFF Patient Registry) had longitudinal PFT measures available.

**Figure 4:**
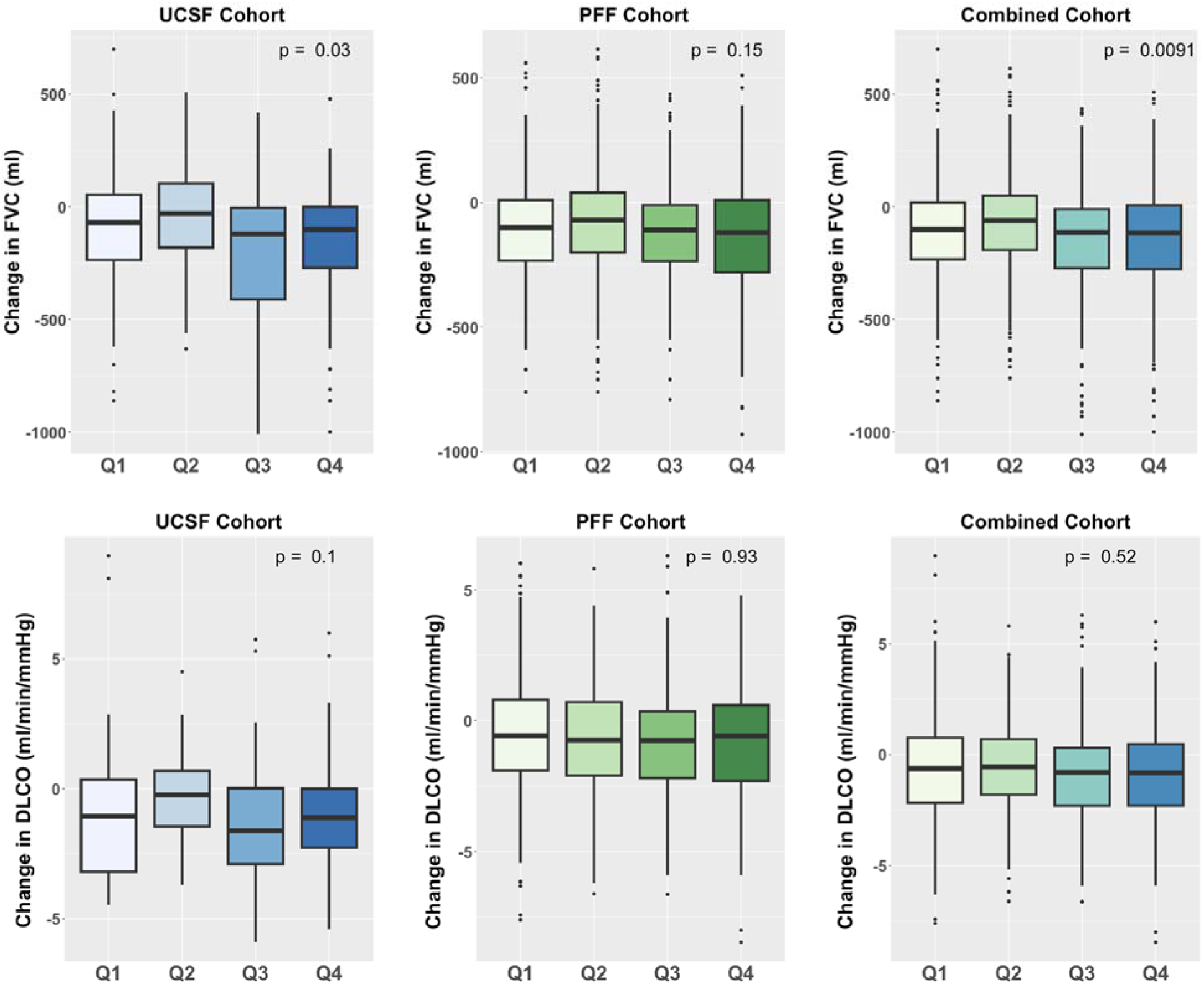
Serum CTHRC1 levels are associated with 1-year changes in lung function Association of serum CTHRC1 levels in IPF with lung disease progression over 1 year by patient cohort. Top: Comparison of 1-year change in FVC (ml) between quartiles of serum CTHRC1 level in UCSF (left), PFF (middle), and combined (right) cohorts. Bottom: Comparison of 1-year change in DLCO between quartiles among cohorts.

Higher CTHRC1 levels were associated with a greater decline in FVC (ml) in the UCSF cohort (mean change in ml per year Q1 = -93, Q2 = -40, Q3 = -206, Q4 = -175, p=0.03) and combined cohort (Q1 = -101, Q2 = -71, Q3 = -139, Q4 = -145, p=0.009) and demonstrated a trend in the PFF Patient Registry (Q1 = -103, Q2 = -79, Q3 = -118, Q4 = -132, p=0.15). Although there was a trend present in the UCSF cohort, elevated CTHRC1 was not associated with a greater decline in DLCO% predicted (p > 0.1).

### CTHRC1 is associated with worse transplant-free survival

In both UCSF and PFF Patient Registry, CTHRC1 level was associated with survival with higher CTHRC1 levels predicting worse mortality, Figure 5. In the UCSF cohort, IPF patients in the quartile with the highest levels of CTHRC1 (Q4) had a significantly worse 5-year transplant-free survival compared to patients with the lower CTHRC1 levels (p = 0.03). This association was more apparent in the PFF cohort, where each quartile increase in CTHRC1 level was associated with lower transplant-free survival (p=<0.001). In the combined cohort after adjusting for age, sex, smoking and baseline FVC%, hazard ratio of a terminal event with the highest levels of CTHRC1 (Q4) was 1.31 (95% CI 1.05-1.91) using patients with the lowest levels of CTHRC1 (Q1) as reference.

**Figure 5:**
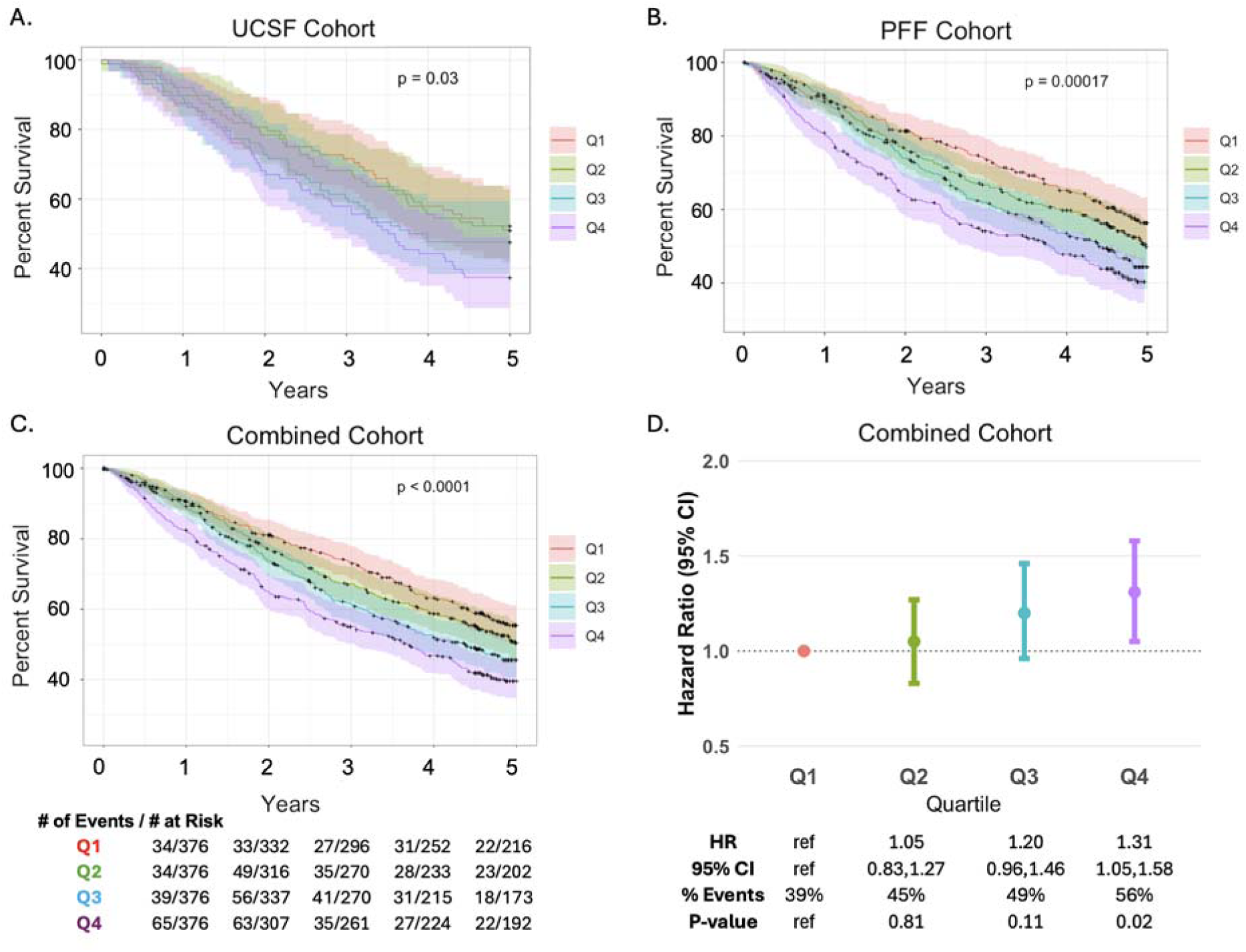
Serum CTHRC1 levels are associated with 5-year transplant-free survival in IPF patients Kaplan Meier survival plot depicting survival by CTHRC1 quartiles for (A) UCSF cohort, (B) PFF and (C) combined cohort with number of events and total number at risk per year. (D) Hazard ratio (HR) of the association between CTHRC1 serum quartile and transplant-free survival adjusted for age, sex, smoking and baseline FVC%. CI, confidence interval.

### Serum CTHRC1 levels increase over time and associate with organ level expression

Serial serum CTHRC1 levels were measured in 39 IPF patients with a mean interval time of 2.7 ± 1.7 years between serum samples. The mean rate of change was an increase of 2288±4211 pg/ml per year. Due to the limited sample size, associations between rate of change and clinical variables could not be robustly assessed. There was a trend toward increased rate of change among patients requiring lung transplantations compared to those who did not (3268±5087 vs. 1357±3014 pg/ml/yr, p=0.15 (Supp 3).

Among the 39 IPF patients, 18 subjects underwent lung transplantation and had tissue available for analysis of CTHRC1 expression at the organ level along with 10 healthy control lungs. CTHRC1 expression was significantly elevated in IPF lungs when compared to controls, p <0.001 (Supp 4a). When serum CTHRC1 concentrations were compared with lung CTHRC1 expression, an overall positive trend of higher CTHRC1 expression in lung tissue with higher serum levels at baseline (p=0.06, Supp 4b) and at time of transplant (p=0.08, Supp 4c).

## Discussion

This study investigated the association of serum CTHRC1, a marker of a novel pathologic fibroblast, and IPF disease severity in two large cohorts of IPF patients. We found higher CTHRC1 serum levels were associated with the presence of IPF, increased disease severity at baseline, disease progression, and overall reduced 5-year transplant-free survival. In addition to the role of CTHRC1 as a potential biomarker in IPF, these findings further support the pathologic role of CTHRC1+ fibroblasts in lung fibrosis.

CTHRC1 was first described in the context of vascular injury, where its expression was found to be transiently upregulated in fibroblasts and smooth muscle cells of balloon injury-induced arteries in rat models.^12^ In human vasculature, CTHRC1 was found in the matrix of calcified atherosclerotic plaques and notably absent in normal arteries. These studies and others showed CTHRC1 expression increased in response to transforming growth factor beta (TGFB) and CTHRC1 overexpression in fibroblasts was associated with increased migration capacity.^13,23^ In murine models, the addition of exogenous CTHRC1 has been shown to upregulate fibroblast proliferation and promote differentiation into activated fibroblasts, leading to increased ECM deposition.^24^ Since its initial discovery, CTHRC1 has been implicated in conditions of aberrant wound healing and tissue repair including myocardial infarction, lung and liver fibrosis, and rheumatoid arthritis.^15,16,25,26^

There is an evolving understanding of the role of CTHRC1 in lung fibrosis. In IPF, CTHRC1 was first described as part of a novel bulk lung gene signature that delineates IPF patients from control subjects and is associated with disease severity.^18^ Interestingly, that study also showed pirfenidone attenuated CTHRC1 expression in the murine bleomycin lung fibrosis model and primary human lung fibroblasts. ScRNA-seq has enabled further characterization of CTHRC1 expression and its role in the lung. While defining the spectrum of fibroblast heterogeneity, a subset of fibroblasts characterized by high CTHRC1 expression was uniquely present in the lungs of mice with fibrosis mediated by bleomycin and in patients with IPF and scleroderma.^10^ These CTHRC1 fibroblasts express the highest level of collagens and ECM components including COL1A1, COL3A1, and POSTN and are localized to fibroblast foci, sites of collagen production and fibroblast proliferation. Ablation of CTHRC1 expressing fibroblasts using a CTHRC1-CreER mouse model caused a significant reduction in lung hydroxyproline content following bleomycin treatment. Lineage tracing demonstrates that CTHRC1 fibroblasts originate from alveolar fibroblasts following lung injury and emerge in response to TGFB, suggesting these fibroblasts are a major source of pathologic matrix production in IPF lungs.^11^ However, global knockout of CTHRC1 is actually associated with increased pulmonary fibrosis in mice injured with intratracheal bleomycin,^27^ suggesting that, while increased CTHRC1 expression is a feature of pathologic fibroblasts, CTHRC1 might be upregulated as a homeostatic mechanism to limit the magnitude of fibrosis.

Our findings suggest that circulating CTHRC1 levels could serve as a useful biomarker in patients with IPF. We show that CTHRC1 serum level is significantly elevated in IPF patients and associated with both disease severity at the time of CTHRC1 measurement and disease course. Notably, the degree of CTHRC1 elevation is associated with transplant-free survival in a dose-dependent manner. While limited in sample size, we also show an association between serum CTHRC1 level and CTHRC1 expression in the lung and that serum CTHRC1 increases over time. Therefore, it can be hypothesized that circulating CTHRC1 levels may represent the burden of pathologic fibroblasts in the lung and thus the burden of active fibrogenesis and eventual lung scarring.

With the advent of single-cell technologies, pathologic expression of numerous markers has been identified at the single cell level in IPF lungs. However, the clinical significance and utility of these markers often remain unclear and untested. Our prior work has established the relevance of CTHRC1 to lung fibrosis, highlighting it as a promising biomarker candidate. Prior to our study, CTHRC1 gene expression in the lung was shown to be associated with disease severity as part of a bulk lung gene signature in IPF. However, due to the difficulty of obtaining tissue for RNA analysis, this may not have practical application in the clinical setting. Here, we measured CTHRC1 using a commercially available ELISA using accessible patient serum, allowing for a non-invasive method for assessing disease severity and progression feasible for clinical use. Our findings demonstrate the utility of leveraging granular omics data from tissue to identify and validate promising molecular markers of disease activity. The application of serum CTHRC1 levels in the clinic would require confirmation of these findings using a CLIA approved platform.

There are both strengths and limitations to this study. Strengths include our ability to examine a novel biologically relevant biomarker in two large, well-characterized, cohorts of IPF patients from multiple centers. This study, therefore, contributes to the overall understanding of IPF pathogenesis while providing evidence for a clinically actionable biomarker associated with a specific subtype of pathologic fibroblast. We demonstrated CTHRC1 expression was predominantly expressed in fibroblasts in lung tissue, however we recognize there may be other organ sources for circulating CTHRC1. The fact that serum CTHRC1 levels are significantly elevated in the IPF cohort compared to healthy controls suggests that the incremental increase in circulating CTHRC1 in IPF is likely coming from the pathologic lungs. Importantly, CTHRC1 was measured both at the tissue level and longitudinally in the serum, providing initial insights into the relationship between circulating and organ-specific expression. However, the sample size of these analyses was limited, restricting our ability to draw definitive conclusions on temporal changes. Therefore, further work needs to be done to confirm the trajectory of CTHRC1 over time and to examine how effective therapeutic interventions longitudinally affect CTHRC1 levels in individual patients.

In conclusion, circulating CTHRC1 levels are elevated in patients with IPF and associated with disease severity and overall survival. The degree of CTHRC1 elevation is strongly associated with survival, suggesting that serum CTHRC1 levels may reflect the load of pathologic fibroblasts present in the lung. Collectively, these findings further contribute to the biologic significance of CTHRC1 in IPF as a pathologic marker of lung fibrosis that may serve as a prognostic biomarker in IPF.

## Data Availability

All data produced in the present study are available upon reasonable request to the authors

## Acknowledgements

We thank all the patients who participated in the UCSF and PFF patient registry. We also thank investigators and other staff at participating PFF Care Centers for providing clinical data, the PFF which established and has maintained the Patient Registry since 2016, and lastly, the many generous donors. The work was supported by AbbVie, Rheumatology Research Foundation, and Nina Ireland Program for Lung Health.

## References

1. Raghu G, Remy-Jardin M, Myers JL, et al. Diagnosis of Idiopathic Pulmonary Fibrosis. An Official ATS/ERS/JRS/ALAT Clinical Practice Guideline. Am J Respir Crit Care Med. 2018;198(5):e44–e68. doi:10.1164/rccm.201807-1255ST

2. Nathan SD, Shlobin OA, Weir N, et al. Long-term course and prognosis of idiopathic pulmonary fibrosis in the new millennium. Chest. 2011;140(1):221–229. doi:10.1378/chest.10-2572

3. Ley B, Ryerson CJ, Vittinghoff E, et al. A multidimensional index and staging system for idiopathic pulmonary fibrosis. Ann Intern Med. 2012;156(10):684–691. doi:10.7326/0003-4819-156-10-201205150-00004

4. Ley B, Collard HR, King TE. Clinical course and prediction of survival in idiopathic pulmonary fibrosis. Am J Respir Crit Care Med. 2011;183(4):431–440. doi:10.1164/rccm.201006-0894CI

5. Wolters PJ, Collard HR, Jones KD. Pathogenesis of idiopathic pulmonary fibrosis. Annu Rev Pathol. 2014;9:157–179. doi:10.1146/annurev-pathol-012513-104706

6. Pardo A, Selman M. The Interplay of the Genetic Architecture, Aging, and Environmental Factors in the Pathogenesis of Idiopathic Pulmonary Fibrosis. Am J Respir Cell Mol Biol. 2021;64(2):163–172. doi:10.1165/rcmb.2020-0373PS

7. Adams TS, Schupp JC, Poli S, et al. Single-cell RNA-seq reveals ectopic and aberrant lung-resident cell populations in idiopathic pulmonary fibrosis. Sci Adv. 2020;6(28):eaba1983. doi:10.1126/sciadv.aba1983

8. Habermann AC, Gutierrez AJ, Bui LT, et al. Single-cell RNA sequencing reveals profibrotic roles of distinct epithelial and mesenchymal lineages in pulmonary fibrosis. Sci Adv. 2020;6(28):eaba1972. doi:10.1126/sciadv.aba1972

9. Reyfman PA, Walter JM, Joshi N, et al. Single-Cell Transcriptomic Analysis of Human Lung Provides Insights into the Pathobiology of Pulmonary Fibrosis. Am J Respir Crit Care Med. 2019;199(12):1517–1536. doi:10.1164/rccm.201712-2410OC

10. Tsukui T, Sun KH, Wetter JB, et al. Collagen-producing lung cell atlas identifies multiple subsets with distinct localization and relevance to fibrosis. Nat Commun. 2020;11(1):1920. doi:10.1038/s41467-020-15647-5

11. Tsukui T, Wolters PJ, Sheppard D. Alveolar fibroblast lineage orchestrates lung inflammation and fibrosis. Nature. 2024;631(8021):627–634. doi:10.1038/s41586-024-07660-1

12. Pyagay P, Heroult M, Wang Q, et al. Collagen triple helix repeat containing 1, a novel secreted protein in injured and diseased arteries, inhibits collagen expression and promotes cell migration. Circ Res. 2005;96(2):261–268. doi:10.1161/01.RES.0000154262.07264.12

13. LeClair RJ, Durmus T, Wang Q, Pyagay P, Terzic A, Lindner V. Cthrc1 Is a Novel Inhibitor of Transforming Growth Factor-β Signaling and Neointimal Lesion Formation. Circ Res. 2007;100(6):826–833. doi:10.1161/01.RES.0000260806.99307.72

14. Yamamoto S, Nishimura O, Misaki K, et al. Cthrc1 Selectively Activates the Planar Cell Polarity Pathway of Wnt Signaling by Stabilizing the Wnt-Receptor Complex. Dev Cell. 2008;15(1):23–36. doi:10.1016/j.devcel.2008.05.007

15. Myngbay A, Bexeitov Y, Adilbayeva A, et al. CTHRC1: A New Candidate Biomarker for Improved Rheumatoid Arthritis Diagnosis. Front Immunol. 2019;10:1353. doi:10.3389/fimmu.2019.01353

16. Li J, Wang Y, Ma M, et al. Autocrine CTHRC1 activates hepatic stellate cells and promotes liver fibrosis by activating TGF-β signaling. EBioMedicine. 2019;40:43–55. doi:10.1016/j.ebiom.2019.01.009

17. Sial N, Ahmad M, Hussain MS, et al. CTHRC1 expression is a novel shared diagnostic and prognostic biomarker of survival in six different human cancer subtypes. Sci Rep. 2021;11(1):19873. doi:10.1038/s41598-021-99321-w

18. Bauer Y, Tedrow J, de Bernard S, et al. A novel genomic signature with translational significance for human idiopathic pulmonary fibrosis. Am J Respir Cell Mol Biol. 2015;52(2):217–231. doi:10.1165/rcmb.2013-0310OC

19. Satija R, Farrell JA, Gennert D, Schier AF, Regev A. Spatial reconstruction of single-cell gene expression data. Nat Biotechnol. 2015;33(5):495–502. doi:10.1038/nbt.3192

20. Samuel Marsh, Maëlle Salmon, Paul Hoffman. samuel-marsh/scCustomize: Version 2.1.2. Published online February 28, 2024. doi:10.5281/ZENODO.5706430

21. Wang BR, Edwards R, Freiheit EA, et al. The Pulmonary Fibrosis Foundation Patient Registry. Rationale, Design, and Methods. Ann Am Thorac Soc. 2020;17(12):1620–1628. doi:10.1513/AnnalsATS.202001-035SD

22. Raghu G, Collard HR, Egan JJ, et al. An official ATS/ERS/JRS/ALAT statement: idiopathic pulmonary fibrosis: evidence-based guidelines for diagnosis and management. Am J Respir Crit Care Med. 2011;183(6):788–824. doi:10.1164/rccm.2009-040GL

23. LeClair R, Lindner V. The role of collagen triple helix repeat containing 1 in injured arteries, collagen expression, and transforming growth factor beta signaling. Trends Cardiovasc Med. 2007;17(6):202–205. doi:10.1016/j.tcm.2007.05.004

24. Duan X, Yuan X, Yao B, et al. The role of CTHRC1 in promotion of cutaneous wound healing. Signal Transduct Target Ther. 2022;7(1):183. doi:10.1038/s41392-022-01008-9

25. Ruiz-Villalba A, Romero JP, Hernández SC, et al. Single-Cell RNA Sequencing Analysis Reveals a Crucial Role for CTHRC1 (Collagen Triple Helix Repeat Containing 1) Cardiac Fibroblasts After Myocardial Infarction. Circulation. 2020;142(19):1831–1847. doi:10.1161/CIRCULATIONAHA.119.044557

26. Peters A, Jyothula S, Huang H, Karmouty-Quintana H. Transitional stem cells and pulmonary fibrosis in non-reversible COVID-19 requiring lung transplantation. In: 12.02 *- ILD/DPLD of Known Origin*. European Respiratory Society; 2023:5. doi:10.1183/23120541.LSC-2023.5

27. Binks AP, Beyer M, Miller R, LeClair RJ. Cthrc1 lowers pulmonary collagen associated with bleomycin-induced fibrosis and protects lung function. Physiol Rep. 2017;5(5):e13115. doi:10.14814/phy2.13115

