## Supplementary material for "Circulating CTHRC1 Levels are Associated with IPF Disease Severity and Survival": Supp 1

**Supplementary Fig 1**

**qPCR Primers**

hCTHRC1:

 Forward:            5’-CCAAGGGGAAGCAAAAGG-3’

Reverse:              5’-CCCTTGTAAGCACATTCCATTA-3’

hCOL1A1 :

Forward:             5’-GGGATTCCCTGGACCTAAAG-3’

Reverse:              5’-GGAACACCTCGCTCTCCA-3’

hGAPDH :

Forward:             5’-AGCCACATCGCTCAGACAC-3’

Reverse:              5’-GCCCAATACGACCAAATCC-3’
