## Supplementary material for "Circulating CTHRC1 Levels are Associated with IPF Disease Severity and Survival": Supp 2

**Supplementary Fig 2**

| **Comorbidities in UCSF cohort** | **Frequency N (%)** |
| --- | --- |
| Gastroesophageal Reflux Disease (GERD) | 130 (37) |
| Coronary Artery Disease (CAD) | 84 (24) |
| Chronic Obstructive Pulmonary Disease (COPD) | 78 (22) |
| Obstructive Sleep Apnea | 77(22) |
| Asthma | 50 (14) |
| Heart Failure | 34 (10) |
| Pulmonary Hypertension | 17 (5) |
