## Supplementary figures and images for "Circulating CTHRC1 Levels are Associated with IPF Disease Severity and Survival"

### Supp 3

**Supplemental Figure 3**

**CTHRC1 rate of change by transplant**

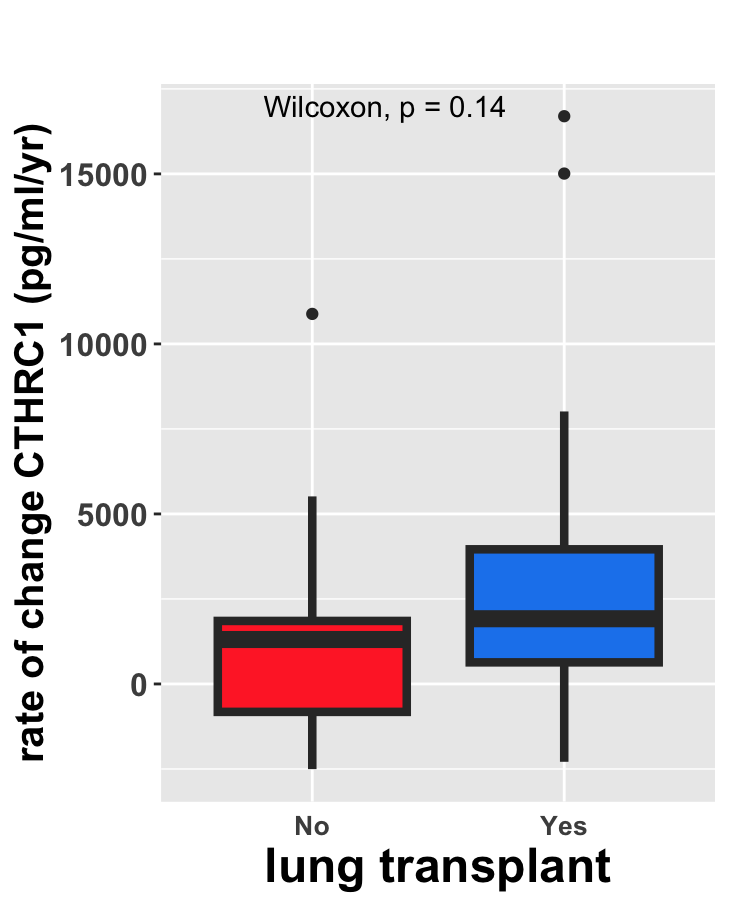
