## Supplementary material for "Circulating CTHRC1 Levels are Associated with IPF Disease Severity and Survival": Supp 4

**Supplemental Figure 4**

**a.**

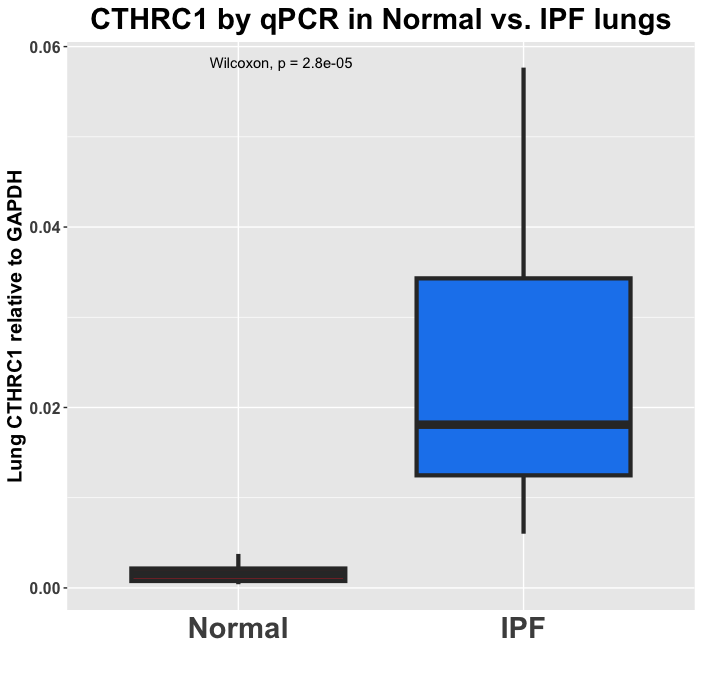

**b.**

Lung CTHRC1 vs. Baseline Serum CTHRC1

Baseline Serum CTHRC1 pg/ml

| Lung CTHRC1 relative to GAPDH | CTHRC1S |
| --- | --- |
| 0.0139304 | 27124.6 |
| 0.0337019 | 46081.1 |
| 0.0361367 | 26567.3 |
| 0.0204399 | 23831.8 |
| 0.0143219 | 31075.9 |
| 0.0376752 | 21450.9 |
| 0.00601253 | 8887.67 |
| 0.0128807 | 45513.4 |
| 0.0157549 | 37813.4 |
| 0.0300407 | 47235.8 |
| 0.0215623 | 27732.4 |
| 0.00631667 | 29404.2 |
| 0.00835069 | 17296.9 |
| 0.0112759 | 28391 |
| 0.0371997 | 70031.9 |

**c.**

Lung CTHRC1 vs. Transplant Serum CTHRC1

Transplant Serum CTHRC1 pg/ml

| Lung CTHRC1 relative to GAPDH | Serum |
| --- | --- |
| 0.0139304 | 35635.1 |
| 0.0337019 | 52687 |
| 0.0361367 | 31623.2 |
| 0.0204399 | 33750.8 |
| 0.0143219 | 47438.4 |
| 0.0376752 | 34946.3 |
| 0.00601253 | 29302.8 |
| 0.0128807 | 43933.1 |
| 0.0157549 | 27479.2 |
| 0.0300407 | 49931.1 |
| 0.0215623 | 28796.3 |
| 0.00631667 | 30062.7 |
| 0.00835069 | 26567.3 |
| 0.0112759 | 47843.7 |
| 0.0371997 | 71582.5 |
